# Peruvian National Survey of Mental Health and Service Utilization in the third year of the COVID-19 pandemic: Protocol for a nationally representative multistage survey

**DOI:** 10.1101/2023.02.22.23286197

**Authors:** Victor Cruz, Andres Pariamachi, Joana Napanga, Brian Pena, Lisette Gamboa, Caroline Gonzales, Paula Delgado, Paula A Burela, Julio Villa-Palomino, Liz Valentin, Delia Zuniga, Orlando Quispe, Pedro Lopez, Jessica Alcantara, Nella Bonilla, Sandra Anton, Mirella Gutierrez, Carlos Diaz, Juan Claux, Alberto Gonzales, Roxana Vivar, Gloria Lobe, Erika Contreras, Blanca Mahr, Milagros Pampamallco, Silvia Salazar, Merle Santos, Veronica Valentin, Jose P Arias, Militza Alvarez, Cindy Amaro, Karla Calderon, Jose Canchis, Fanny Carbajal, Jessica Chire, Liany Correa, Linnette Hermoza, Giannina Moron, Candy Palomino, Rocio Ramirez, Edelmira Rojas, Mercedes Arevalo, Cesar Arellano, Vanessa Herrera, Janet Ricardi, Gloria Gupio, Onesimo Jaramillo, Carmen Clapes, Esther Cerna, Mariluz Antunez, Fernando Luna, Alfredo Saavedra, Sara Carbajal, Favio Vega, Paulo Ruiz, Javier Saavedra, Elba Luna, Javier del Campo, Itziar Familiar-Lopez, Amantia A Ametaj, William E Copeland, Jim Anthony, Rafael Nishimura, Henning Tiemeier, Bizu Gelaye

## Abstract

**Background:** Peru is the worst affected country by the COVID-19 pandemic showing the world highest mortality rate, thus triggering an increased mental health burden. Nevertheless, there are few population-based epidemiologic surveys of mental disorders in Peru; Therefore, nationally representative research is needed to understand the underlying population-based mental health burden and identify unmet care needs.

**Objective:** The present study aims to estimate the prevalence and patterns of psychiatric disorders, mental health service use, and unmet mental health care needs

**Methods:** This cross-sectional study will collect information from a multistage random sample of 19,500 households. A child, an adolescent, an adult, and an older adult will be interviewed in the household. Trained staff will conduct face-to-face diagnostic interviews via the Preschool Age Psychiatric Assessment, the Child and Adolescent Psychiatric Assessment, the WHO’s Composite International Diagnostic Interview, and the Alzheimer Disease 8 Scale. In addition, descriptive and inferential analysis for complex sampling will be performed to estimate the prevalence and correlates.

**Ethics and dissemination:** IRB will approve the research protocol before the commencement of the study. Only respondents who signed their informed consents or assents will participate in the study; The parent or guardian will sign the consent for the participation of preschool children. The research findings will be disseminated in peer-reviewed publications, scientific reports, and presentations at national, and international meetings. In addition, de-identified data and study results will be posted on the Peruvian National Institute of Mental Health (PNIMH) website to be freely available to policymakers, researchers, and the general public.

**Strengths and Limitations:**

- This will be the first national survey on mental health and services use with a large probabilistic sample size, allowing to estimate the prevalence of psychiatric disorders and service use for the rural and urban areas of each of the 25 Peruvian regions.
- At the national level, this survey will have enough power to estimate the prevalence of rare psychiatric disorders with a prevalence closest to one percent.
- Researchers aim to conduct a second interview among the participants after at least one year of performing the first evaluation to estimate the incidence of psychiatric disorders.
- Researchers are advocating among the Peruvian Ministry of Economics and Finance officers to conduct a national survey each year to evaluate the impact of mental health policies via the analysis of trends.
- The principal limitation of this study is its cross-sectional design which does not allow to infer the temporality of associations.

## INTRODUCTION

The prevalence of mental and substance use disorders increased by 37.6% between 1990 and 2010, becoming one of the significant contributors to the burden of disease worldwide (1). Furthermore, more than 80% of the people affected reside in low-and middle-income countries (LAMICs), creating substantial social (2–4) and economic hardships (5–8) for affected individuals and their families and society as a whole (9,10). For this reason, several organizations, including the World Health Organization (WHO), advocate primary prevention, control, and treatment of mental health disorders and psychosocial problems, especially those starting in the early years of life (11).

Peru has been experiencing an epidemiologic transition in the last few decades due to its fast economic development, urbanization, and decreasing fertility and mortality rate (12,13). Consequently, chronic non-communicable diseases (NCDs) are now more significant contributors to disease burden than infectious diseases, especially major depression and alcohol use disorders (14). Psychosocial problems such as intimate partner violence (IPV) and child abuse (CA), often understudy in most mental health surveys, affect disproportionally LAMICs, such as Peru (15,16). Despite their enormous burden, mental health disorders and psychosocial problems remain under-recognized and undertreated. In Peru, as in most LAMICs, large psychiatric hospitals remain the dominant facilities of the mental health care system today. According to estimates from 2012, one in five Peruvians had a mental health condition, and only one in five of those people received the care they needed. Sadly, like for other health conditions in Peru, this pattern is not homogenous across groups. Those with economic hardships and victims of historical trauma and political violence are disproportionally affected (17).

Integrating mental health services in the community and primary health care settings has been one of the strategies suggested by the WHO and other organizations to adequately address the burden of mental disorders. In the last five years, the Peruvian Ministry of Health has been implementing a community mental health service reform to address the epidemiologic transition of chronic NCDs and the absence of mental service outside Lima, the capital city of Peru (18,19). However, mental health disparities remain very high, especially in rural areas where the unsatisfied necessity of mental health services reaches 90% of the population with psychiatric disorders, while in the urban areas is around 70% (20). In response to this need in 2019, Peru passed a new national mental health law that increased community mental health centers (CMHCs) (21). The CMHCs bring mental health services from psychiatric hospitals to local settings, where providers engage patients and communities as partners. Peru has the highest excess mortality rate due to the COVID outbreak in the world, with a rate of 590 per 100,000 inhabitants (22). The COVID-19 pandemic presented countries such as Peru with significant economic, political, scientific, and public health challenges and long-lasting impacts (23). The high COVID morbidity/mortality rate and the public health measures, which were essential interventions to limit the transmission of Covid-19, came with adverse population mental health consequences and behavioral outcomes, including major depression, anxiety disorders, stigmatization attitudes from others, posttraumatic stress disorder, stress, acute and chronic sleep disturbances, and increases use of alcohol and tobacco (24–28). While the pandemic brought the importance of mental health to the general public and policymakers, the extent of mental health problems and unmet needs remains unknown.

National mental health surveys are critical for health policy planning (29), especially for studying the underlying burden of disease, mental health services use, treatment access, and existing barriers (30– 32) as well as for mental health promotion and prevention (33). Therefore we designed the present study to address the following aims: (1) to estimate prevalence and patterns of psychiatric disorders, alcohol, tobacco, and illegal substance use and abuse in a nationally representative population; (2) to estimate the prevalence of the principal psychosocial problems such as intimate partner violence and child abuse; (3) to reliably estimate the 12-months proportion of mental health service demand by disorder; (4) to estimate the lifetime and 12-months use or access of mental health services by disorder, professional type and service sector; (5) To estimate the proportion of perceived satisfaction and helpfulness of mental health treatment received by disorder and professional type; (6) to estimate the 12-months proportion of unmet demand of mental health services by disorder; (7) to estimate the population patterns of perceived barriers for delaying the access to mental health services; (8) identify the patterns of mental health care seeking and service utilization; and (9) identify groups at high risk for substance use disorder so that mental health care services and programs can be developed.

## METHODS

### Study Design

The design of the present survey research is a cross-sectional study with a national probabilistic sample of 19,500 dwelling units where an adult (18-59 years old), an adolescent (9-17 years old), a child (2-8 years old) and an older adult (60+ years old) will be interviewed. The exclusion criteria will be hearing impairment, cognitive or neurologic conditions, and difficult verbal communication. The survey will be delivered through house-to-house interviews using a computer-assisted interview.

### Study setting, sampling strategy, and participants

The study population of the present survey will be the residents of the 25 Peruvian regions (Figure 1). From October 2022 to January 2023, the data will be collected via a stratified multistage probabilistic household sampling. The strata will be each of the 25 Peruvian regions and the rural and urban areas inside each region. Moreover, the sampling strategy will consist of three stages. In the first stage, census clusters will be selected via a simple random process. Next, the interviewers will prepare a register with all the dwelling units belonging to the selected census cluster; this register will be used in the second sampling stage to choose dwelling units. In the second stage, a dwelling unit will be selected randomly inside the cluster via a systematic random selection process; in these selected units, the interviewers will register all the potential participants inside the selected dwelling units who fulfill the research inclusion criteria. They will only interview the participants who agree to participate and sign a written informed consent. Parental consent will be garnered prior to participation for those younger than 18. In the third stage, a designed respondent will be randomly selected inside each house using the Kish table. The number of respondents will be proportional to the population size of each region and, within each region, proportional to the urban and rural population size.

Previously trained interviewers, university students of psychology, midwifery, and nursing who get their bachelor’s degrees, will employ face-to-face computer-assisted interviews using the Spanish version of the psychiatric diagnostic interviews.

**Figure 1.**
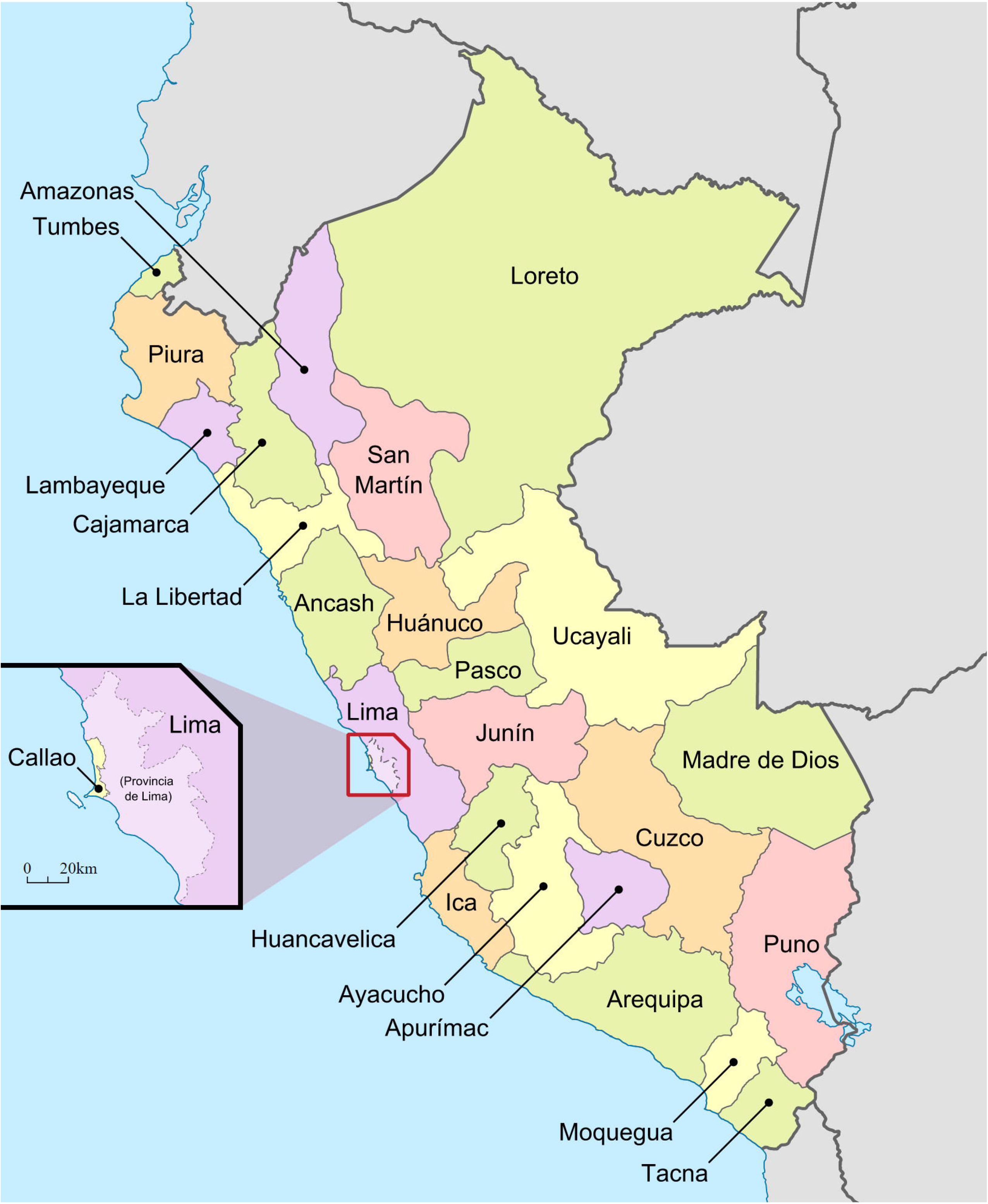
Peruvian political map with its 25 regions.

Source: https://www.mapsland.com/south-america/peru/large-detailed-administrative-map-of-peru

### Sample Size

The target sample size was determined assuming a 12-month prevalence (p) for depression, one of the leading causes of disease burden in Peru would be 12%, and a design effect of 1.5. The following formula was used to achieve a coefficient of variation (CV) of 2% (34): n=(1-p)/(CV^2*p+(1-p)/N), which would require a sample size of roughly 19,500 households. Assuming an eligibility rate of 96% and a response rate of 70%, this would require a sample size of 28,785 households.

### Study Procedures

The data will be gathered using standardized diagnostic assessments via the Preschool Age Psychiatric Assessment (PAPA) for children aged 2 to 8 years (35). For adolescents aged 9 to 17 years, the Child and Adolescent Psychiatric Assessment (CAPA) will be used (Angold et al., 1995), and the WHO Composite International Diagnostic Interview (CIDI) will be used for people 18 to 59 years (36). In addition, all these instruments will be translated into computer-assisted DSM-5 version (37).

Additional data will be gathered using cross-culturally validated screening instruments. Depressive symptoms will be measured with the Patient Health Questionnaire-9 (PHQ-9) (38), previously validated in Peru (39). The anxiety symptoms will be measured by the General Anxiety Disorder-7 (GAD-7) (40).

For older adults (60 years and older), Washington University’s Dementia screening test (AD-8) and the Mini-Cog will be administered. The AD-8 is a brief instrument used to help discriminate between signs of normal aging and mild dementia (41). The Mini-Cog is a brief (3-item) recall test of memory and a scored clock drawing test (42). As well as the PHQ-9, the GAD-7, the Columbia–Suicide Severity Rating Scale (C-SSRS) (43), the WHO Alcohol, Smoking and Substance Involvement Screening Test (ASSIST) (44), the Pittsburgh sleep quality index (PSQI) (45), the 12-Item Short-Form Health Survey (12-SFHS) (46), and the UCLA Loneliness Scale version 3 (47).

### Training of Research Staff

Research assistants and interviewers will be trained to administer the computer-assisted version of the PAPA, CAPA, and CIDI. Research assistants will include psychiatrists, psychologists, social workers, anthropologists, and sociologists. Recent university bachelor’s graduates from health-related or social sciences programs will be in charge of recruitment and consenting.

The study investigators will develop training materials on human subjects research procedures, recruitment consenting, and data collection through questionnaires, sampling, and physical measurement protocols, with continuing refresher training provided periodically. In addition, a 3-day training program for PAPA, CAPA, and CIDI will be organized. All research team members will receive human subjects training prior to the commencement of the project. This training has ensured that all team members understand the study’s overall purpose, why special measures are being used, and other ethical research issues, such as respect, beneficence, and confidentiality. The goals of the training will include helping staff develop a basic understanding of mental health, its characteristics and causes, and its impact on the health of men, women, and children; helping them understand the goals of the research project; and helping them become familiar with the questionnaires and the protocol of the survey. All training materials will be available for reference.

In addition, research staff will get training on interviewing skills and following safety and ethical guidelines while conducting the study. Trainees will participate in mock interviews with senior investigators. These role-play sessions will be closely mentored, observed, and supervised.

### Demographics and Mental Health Services

Sociodemographic variables will be evaluated via the sociodemographic modules from the CIDI, CAPA, and PAPA. In addition, mental health service use will be evaluated via the mental health service access module from the CIDI and the Child and Adolescent Service Access (CASA) from the CAPA (48). The following CIDI question will evaluate the 12-month perceived mental health demand: “Was there ever a time during the past 12 months when you felt that you might need to see a professional because of problems with your emotions or nerves or your use of alcohol and drugs?” In addition, the 12-month mental healthcare access by disorder will be evaluated by the CIDI question: “Did you receive professional treatment for your (…) at any time in the past 12 months?”

### Prevalence of Mental health Disorders

The prevalence of mood disorders, psychotic disorders, anxiety disorders, obsessive-compulsive disorders, trauma-related disorders, substance use disorders, eating disorders, impulse control disorders, and personality disorders among adults 18+ years will be measured via the CIDI (36). Among adolescents aged 9 to 17 years, the CAPA will be used to measure the prevalence of mood disorders, psychotic disorders, anxiety disorders, obsessive-compulsive disorders, trauma-related disorders, substance use disorders, eating disorders, somatization disorders, sleep-wake disorders, elimination disorders, tic disorders/trichotillomania, and disruptive behavior disorders (49). For children aged 2 to 8 years, the PAPA will estimate the prevalence of reactive attachment disorders and almost all CAPA disorders (35).

### Adversity or Trauma History

The lifetime and 12-month prevalence of psychological, physical, and sexual IPV will be measured via the WHO Violence Against Women Instrument (VAWI) among cohabiting partners (50). In addition, the history of trauma exposure related to extreme violence and torture provoked by the Shining Path Maoism guerrilla and the Peruvian army will be measured via the Harvard Trauma Questionnaire (HTC) (51). Finally, the history of psychological, physical, and sexual CA during the adult interviewee’s first 18 years of life will be measured via the Adversity Childhood Experiences (ACE) Study Questionnaire (52).

### Pandemic-related Variables

The history of ever diagnosis and treatment of COVID-19 will be evaluated by questions from the Michigan Collaborative for Infant Mental Health Research (53). These questions also evaluate the COVID-19 pandemic-related stress, isolation and how it affects their job. The isolation was measured via the modified version of the UCLA Loneliness Scale (54).

### Predictor Variables

The CIDI modules on family history of psychopathology, childhood family adversity, social networks and support, and stressful life events and difficulties will be used to identify correlates. Furthermore, Scientific literature shows an association between anxiety, depression, and suicide with high altitude (55,56); researchers have postulated that the high altitude hypoxia and inflammation might be the underlying mechanisms of this association (57). The present study gives an extraordinary opportunity to test this association. The large sample size will give enough power to evaluate this association among people living at different altitudes on the Peruvian Andes while controlling for putative confounders of this association.

### Epidemiological Causal Analysis

We will collect data on demographic and lifestyle variables, providing a rich source of covariates that may operate as confounders, mediators, or effect modifiers. For instance, the Peruvian highlanders are usually Quechuas and Aymaras, who have lived in poverty for more than 200 hundred years since their independence from Spain. Census data shows that the percentage of Quechuas and Aymaras grows accordingly with the village’s altitude, and poverty also increases with the altitude of the villages (58). Furthermore, previous studies demonstrated an association of poverty with depression (59–63), especially when the measure is done with more sensible instruments such as the multidimensional poverty measurement (64). Therefore, the moderation effect of poverty on the relationship between depression and high altitude will be tested in the present study.

### Data Analysis Plan

The survey proportion of the software Stata will be used to estimate the prevalence of psychiatric disorders, service use, and psychosocial problems, such as IPV and CA. The analysis will consider the sample weights and the survey complex design to estimate the Taylor series variance (65). The latent class analysis for complex surveys of the software Mplus will be used to estimate the population patterns of psychiatric disorders at the population level (66). The discrete survival time analysis will be used to estimate the age-specific risk of psychiatric disorders and psychosocial problems across high altitudes (67). For this analysis, the participants’ age will be used as a marker of cohort membership, and cumulative incidence or lifetime prevalence of psychiatric disorders age by age will be estimated for each cohort (68). This procedure will make it possible to trace an event’s occurrence over time without longitudinal data (69,70). Finally, the survey logistic regression will be used to estimate the correlation of high altitude with psychiatric disorders or psychosocial problems, considering the mediator, moderator, and confounding effect of this association (71).

### Patient and Public Involvement

There will not be a formal patient or public advisory committee. Notwithstanding, the participants and the general public will be permanently informed that they can follow everything related to the present survey on the official webpage of the study.

### Ethics and Dissemination

The Institutional Review Board (IRB) from the PNIMH will approve the present research protocol. Only respondents who understand and provide signed informed consent will participate in this study. To be enrolled in this research, people under 18 years will provide informed assent, and their parents must also provide informed consent. Participants can abandon the study if they want to at any interview moment. All participants of this study are assured during the recruitment and consenting process that participation in the project is voluntary. Respondents with a panic attack or suicide ideation will be made sure they receive treatment in the PNIMH or the nearest CMHCs. The data will be stored in the database of the REDcap software at the PNIMH; only de-identified data will be provided to the researchers worldwide for statistical analysis via the webpage of the PNIMH.

The findings of the present survey study will be disseminated via the website of the PNIMH and through peer-reviewed journals. In addition, the study results will be available to the Peruvian national or regional policymakers and laypeople via executive reports and the webpage of the PNIMH.

## DISCUSSION

The present survey study will allow the estimation at the national and regional level of the prevalence of psychiatric disorders, mental health service use, and psychosocial problems such as IPV and CA. In a study carried out in several countries around the world by the WHO, Peru had the highest lifetime prevalence of physical IPV (61%), and Japan had the lowest (13%) (72). The present study has enough power to make accurate estimations of the prevalence of depression and anxiety disorders, the two leading causes of burden of disease in Peru among mental and substance use disorders (73). Furthermore, this study will give more accurate estimates of the association of high altitude with psychiatric disorders, considering the mediational, moderating, and confounding effects.

Training interviewers will gather information via face-to-face interviews at the dwelling unit of the participants of this study. The trained interviewers will use state-of-the-art psychiatric diagnostic interviews based on the DSM-5 and the International Classification of Diseases 10^th^ edition (ICD-10) diagnostic criteria (74). The interviewers, health and social science professionals with bachelor’s degrees, will use the Spanish version of our instruments for Spanish-speaking participants. Moreover, Interviewers fluent in Quechua, Aymara, or Ashaninka will interview participants who only speak these original Peruvian languages. Trained supervisors will follow the survey interviews closely throughout the data-collection process.

Using stratified multistage random sampling and the coefficient of variation (CV) as a measure of precision will allow more accurate estimations in the present survey. The CV is more stringent (gives a bigger sample size) than the absolute or relative errors in calculating the sample size when the prevalence is close to or less than one percent (75). Granting in this way precise estimations of the prevalence of psychiatric disorders for comparisons with the estimates of other mental health epidemiologic studies carried out worldwide, thus contributing to the development of the science of mental health epidemiology following the COVID-19 pandemic. The reliable and valid estimations of the psychiatric disorders prevalence getting from the present survey will fulfill the knowledge gap of the global burden of mental disorders during the COVID-19 pandemic in the southern hemisphere (76)

This survey will be the first to provide national and regional estimates for the 25 Peruvian regions about the prevalence of psychiatric disorders, psychosocial problems, and mental health service use. It will give important information about the burden of mental health disorders and problems and the unmet mental health needs of the Peruvian population. This information will be crucial for planning mental health policies at the regional and national levels. These health policies should look to improve the mental health status of the underserved and vulnerable population, paying due attention to less prevalent disorders such as obsessive-compulsive disorders, eating disorders, or substance use disorders. Nowadays, these disorders lack adequate or specific care programs for these pathologies in the CMHCs that the Peruvian government is implementing as part of its mental health reform plan (17). For instance, eating disorders and substance use disorders have doubled their years lost due to disability (YLD) from 1990 to 2015 in Peru (73).

The principal limitations of the present study are the self-report nature of the survey, which may be subjected to non-systematic errors in recall and systematic non-disclosure errors leading to misclassification. This is an inherent limitation in psychiatric epidemiologic studies. Moreover, cross-sectional studies will not allow for assessing temporal relationships between the onset of mental illness and risk factors, which precludes the determination of causal inference (77).

Due to budget restrictions, the sample size of the present survey was estimated using a coefficient of variation of 2% as a measure of precision and a major depression 12-month prevalence of 12%. In calculating sample size, future Peruvian National Mental Health Surveys (PNMHS) should use a coefficient of variation of 1% and the prevalence of schizophrenia of 0.6%. The lowest prevalence of schizophrenia will increase the sample size of the new surveys allowing accurate estimation of the prevalence of rare psychiatric disorders such as bipolar disorder or obsessive-compulsive disorder (78).

There are few epidemiologic surveys of mental disorders in Peru with up-to-date instruments and rigorous methodology. The PNMHS is critical to identify the frequency and distribution of psychiatric disorders, as well as the treatment and care service gaps, because Peru has been severely affected by the COVID-19 pandemic having the world’s highest excess mortality rate due to COVID-19 (22). Furthermore, worldwide mental disorders and substance use disorders are among the leading cause of the burden of disease, with a clear increased trend in the last decades (79). In Peru, neuropsychiatric diseases are the first cause of disease burden or disability-adjusted life-years (DALYs) (80). Furthermore, in 2019 Peru had the highest amphetamine use disorder (66 DALYs) and opioid use disorder (70 DALYs) burden per 100000 population in South America (81). Therefore, nationally representative research is needed to understand the underlying population-based mental health burden, patterns of mental disorders, and substance use issues and to identify unmet care needs. Such data can be used to (i) create population-based mental health strategies, (ii) map existing resources for the delivery of mental health services, and (iii) develop, implement, evaluate, scale, and sustain meaningful mental health programs and policies supporting the current Peruvian mental health reform of the care services (17).

## Supporting information

STROBE Statement Checklist

## Data Availability

All data produced in the present study are available upon reasonable request to the authors

## Technical Appendix, Statistical Code, and Dataset Available from

The Harvard Dataverse repository, DOI:

## Acknowledgments

The PI of this study wants to especially thank professor James C. Anthony for his valuable insights and teachings in the field of mental health epidemiology.

## Author Contributions

VC, AP, JN, JC, RV, JPA, FL, JS, FV, JA, HT, and BG designed the study, drafted the protocol, and equally contributed to the writing. MG, CD, KC, and RN designed the sample. BP was involved in the adaptation and validation of the instruments. LV training and supervision procedures. All the authors provided critical feedback on the protocol.

## Patient and public involvement

There was no involvement of patients or public advisory committees in this study, but information about the study will be posted on the website of the PNIMH.

## Patient consent for publication

Not applicable.

## Provenance and peer review

Not commissioned; externally peer-reviewed.

## Open access

This will be an open-access article.

